# Breakthrough SARS-CoV-2 infections in MS patients on disease modifying therapies

**DOI:** 10.1101/2022.01.22.22269630

**Authors:** Irene Schiavetti, Cinzia Cordioli, Maria Laura Stromillo, Maria Teresa Ferrò, Alice Laroni, Eleonora Cocco, Gaia Cola, Livia Pasquali, Maria Teresa Rilla, Elisabetta Signoriello, Rosa Iodice, Alessia Di Sapio, Roberta Lanzillo, Francesca Caleri, Pietro Annovazzi, Antonella Conte, Giuseppe Liberatore, Francesca Ruscica, Renato Docimo, Simona Bonavita, Monica Ulivelli, Paola Cavalla, Francesco Patti, Diana Ferraro, Marinella Clerico, Paolo Immovilli, Massimiliano Di Filippo, Marco Salvetti, Maria Pia Sormani, the “Breakthrough infections in MS” study group

## Abstract

**Background:** Patients with Multiple Sclerosis (pwMS) treated with anti-CD20 or fingolimod showed a reduced humoral response to SARS-CoV-2 vaccines. In this study we aimed to monitor the risk of breakthrough SARS-CoV-2 infection in pwMS on different Disease Modifying Therapy (DMT).

**Methods:** Data on number of vaccinated patients and of patients with a breakthrough infection were retrospectively collected in 27 Italian MS centers. We estimated the rate of breakthrough infections and of infection requiring hospitalization per DMT.

**Findings:** 19641 vaccinated pwMS were included in the database. After a median follow-up of 8 months, we observed 137 breakthrough infections. As compared to the other DMTs, the rate of breakthrough infections was significantly higher on ocrelizumab (0.57% vs 2.00%, RR=3.55,95%CI=2.74-4.58, p<0.001) and fingolimod (0.58% vs 1.62%, RR=2.65,95%CI=1.75-4.00, p<0.001), while there were no significant differences in any other DMT group. In the ocrelizumab group the hospitalization rate was 16.7% vs 19.4% in the pre-vaccination era (RR=0.86,p=0.74) and it was 3.9% in all the other DMT groups vs 11.9% in the pre-vaccination period (RR=0.33,p=0.02).

**Interpretation:** The risk of breakthrough SARS-CoV-2 infections is higher in patients treated with ocrelizumab and fingolimod, and the rate of severe infections was significantly reduced in all the DMTs excluding ocrelizumab.

## Introduction

Several recent studies evaluated the effect of vaccination against SARS-CoV-2 in patients with multiple sclerosis (pwMS) treated with disease-modifying therapies (DMTs). There is wide consensus that the use of anti-CD20 monoclonal antibodies and fingolimod are associated with an impaired virus-specific humoral immune response as compared to all the other DMTs^1-4^. On the other hand, there is also growing evidence that vaccinated pwMS treated with anti-CD20 generated robust virus specific CD4 and CD8 T cell responses^4-5^, while these are slightly reduced in fingolimod treated patients^5^. A preliminary follow-up study of 344 fully vaccinated pwMS on DMT reported 13 breakthrough infections, 10 of which were in patients on anti-CD20 therapy and the remaining 3 on fingolimod^6^, suggesting a relevant role of antibodies in preventing the infection. The French registry recently reported a case series of 18 pwMS who had Covid-19 after two doses of BNT162b2-vaccination, 13 of which treated with anti-CD20 and four with fingolimod^7^. Finally, the clinical follow up of the CovXiMS study^1^ evaluating humoral response in 1705 pwMS who received two doses of mRNA vaccines^8^, reported 23 breakthrough infections over a 6 month follow up. The risk of infection was associated with lower SARS-CoV-2 antibody levels measured after 4 weeks from the second vaccine dose^8^.

Against this background and taking advantage of the large network of MS centers within the Italian Alliance against Covid-19 promoted by the Italian MS Society, we collected data from 27 Italian MS centers on the number of vaccinated patients and the number of patients who had a breakthrough infection in each DMT group, in the period preceding the spread of the Omicron variant, that started its massive diffusion in Italy after the December 2021 holiday season. Aim of this study is to estimate the rate of breakthrough infections per DMT class on a large sample of vaccinated pwMS and to compare the rates of severe infections to the rate observed in Italy in the pre-vaccination era^9^.

## Patients and Methods

### Study design and participants

This was a retrospective data collection conducted in 27 Italian MS centers on pwMS undergoing the SARS-CoV-2 vaccination. Each MS center was requested to report the number of pwMS vaccinated by two mRNA vaccine doses (BNT162b2 (Pfizer Inc, and BioNTech), or mRNA-1273 (Moderna Tx, Inc)) in each DMT group from March 2021 to December 25, 2021. Data cutoff was set before the spread of the Omicron variant in Italy, since on December 23, 2021 the percentage of Omicron infections was estimated to be 28% (https://www.iss.it/primo-piano, accessed on December 25, 2021). Breakthrough infections occurred within 8 months, defined as a PCR-confirmed test after 14 days from the second or the third vaccine dose, were extracted from the platform dedicated to Covid-19 data collection in pwMS (MuSC-19 database^10^) for the participating centers. The post-vaccination SARS-CoV-2 infection was recorded in a dedicated Case Report Form (CRF).

The study is done in compliance with the principles of the Declaration of Helsinki. The study was approved by the regional ethics committee of Liguria (University of Genoa; n 130/2020–DB id 10433) and at a national level by the Italian Medicines Agency. Written informed consent was obtained from all participants before starting any study procedures.

### Primary Outcome: breakthrough infection

The primary objective of this analysis was to compare the incidence of breakthrough SARS-CoV-2 infections among the vaccinated pwMS in each DMT group. These conditions entail a PCR-confirmed swab, and a time lag of at least 14 days from a full vaccination cycle (after the second or third vaccination dose, or after the first dose following a Covid-19 infection).

### Statistical analysis

The percentage of patients with a breakthrough infection in the different DMT groups was calculated. 95% Confidence Intervals (CI) were estimated using the normal approximation to the binomial calculation^11^. Difference of rate of infections between DMT groups were estimated by Risk Ratios (RR) and evaluated by Chi-square tests. Difference of rate of infections in the first 4 months vs the second 4 months of follow up were estimated by ORs and evaluated by the McNamar test for paired data.

### Funding

FISM [2021/Special-Multi/001].

## Results

Data were collected between March 1, 2021 and December 24, 2021. 19641 pwMS who had a full vaccination cycle with an mRNA vaccine (2 or 3 vaccine dose, or 1 vaccine dose after Covid-19 infection) were included in the database. The number of vaccinated pwMS in each DMT group is reported in Table 1. The mean follow-up time was 249 days (range 99-354). Among them, 137 breakthrough infections were observed (26 (19.0%) after the third dose, 1 after Covid-19 infection and one dose) over a mean interval after the last vaccine dose of 142 days (range 14-262) (Table 1). Over the whole follow-up of about 8 months, we compared the proportion of patients with breakthrough infections in each DMT group to the pooled proportion of the patients on all the other DMTs (Figure 1, panel A). The rate of breakthrough infections was significantly higher in patients treated with ocrelizumab (2.00%, 95%CI=1.36-2.66) than in patients treated with all the other DMTs (0.57%, 95%CI=0.46%-0.68%) with a RR=3.55, 95%CI=2.74-4.58, p<0.001; the same was observed in patients treated with fingolimod who had a higher rate of breakthrough infections (1.62%, 95%CI= 1.02%-2.21%) than the patients treated with all the other DMTs (0.61%, 95%CI=0.50-0.72) with a RR=2.65, 95%CI= 1.75-4.00, p<0.001. Among the patients who had the SARS-CoV-2 infection, 10 (7.3%) had a severe disease course and were hospitalized. Six patients treated with ocrelizumab were hospitalized and in this group the rate of hospitalization was 16.7%, slightly lower but not significantly different than the pre-vaccination rate observed in Italy (19.4%) in the same DMT group^7^ (relative reduction=14%, RR=0.86, 95%CI=0.38-1.91, p=0.74). In the fingolimod group we observed just 1 hospitalized patient (3.6%). The rate of hospitalization was 3.9% in all the other DMT groups as compared to 11.9% in the pre-vaccination period^7^ (relative reduction=67%, RR=0.33, 95%CI=0.13-0.88, p=0.02). One patient in ocrelizumab was admitted to the Intensive Care Unit (ICU) and recovered.

**Table 1:** Characteristics of patients with breakthrough infections (N = 137)

|  |  |  |
| --- | --- | --- |
| **Female sex** |  | 85 (62.0) |
| **Age, years** |  | 42.3 ± 10.70 |
| **BMI (kg/m2)** |  | 24.6 ± 4.95 |
| **MS phenotype** | Primary progressive | 9 (6.6) |
|  | Relapsing remitting | 116 (84.7) |
|  | Secondary progressive | 7 (5.1) |
|  | Missing data | 5 (3.6) |
| **Disease duration, months** |  | 99.5 (44.0 - 182.0) |
| **Last EDSS before Covid-19 infection** |  | 2.0 (1.0 - 3.5) |
| **Relapse in the six months before Covid-19 infection** |  | 7 (5.1) |
| **Number of breakthrough infections in each DMT/number of vaccinated patients, (%)** | alemtuzumab | 0/371 (0.0) |
|  | azathioprine | 0/298 (0.0) |
|  | cladribine | 4/570 (0.70) |
|  | dimethyl fumarate | 22/2668 (0.82) |
|  | fingolimod | 28/1733 (1.61) |
|  | glatiramer acetate | 4/1514 (0.26) |
|  | interferon | 7/2452 (0.29) |
|  | natalizumab | 15/1843 (0.81) |
|  | ocrelizumab | 36/1794 (2.00) |
|  | rituximab | 3/364 (0.82) |
|  | teriflunomide | 10/1379 (0.73) |
|  | other | 0/389 (0.0) |
|  | untreated | 8/4266 (0.19) |
| **Boost COVID-19 vaccination** |  | 26 (19.0) |
| **Heterologous vaccine** |  | 4 (2.9) |
| **Covid severity** | Asymptomatic, viral RNA detected | 16 (11.8) |
|  | Symptomatic, independent | 95 (69.3) |
|  | Symptomatic, assistance needed | 16 (11.8) |
|  | Hospitalized, no oxygen therapy | 4 (2.9) |
|  | Oxygen by mask or nasal prongs | 5 (3.7) |
|  | Intubation and mechanical ventilation, piO2/FiO2≥150 or SpO2/FiO2≥200 | 1 (0.7) |

*Results are expressed as count (%), mean ± Standard Deviation, or median [Inter Quartile Range], as appropriate.*
*MS=Multiple Sclerosis, BMI=Body Mass Index, EDSS=Expanded Disability Status Scale, DMT=Disease Modifying Therapy*

**Figure 1.**
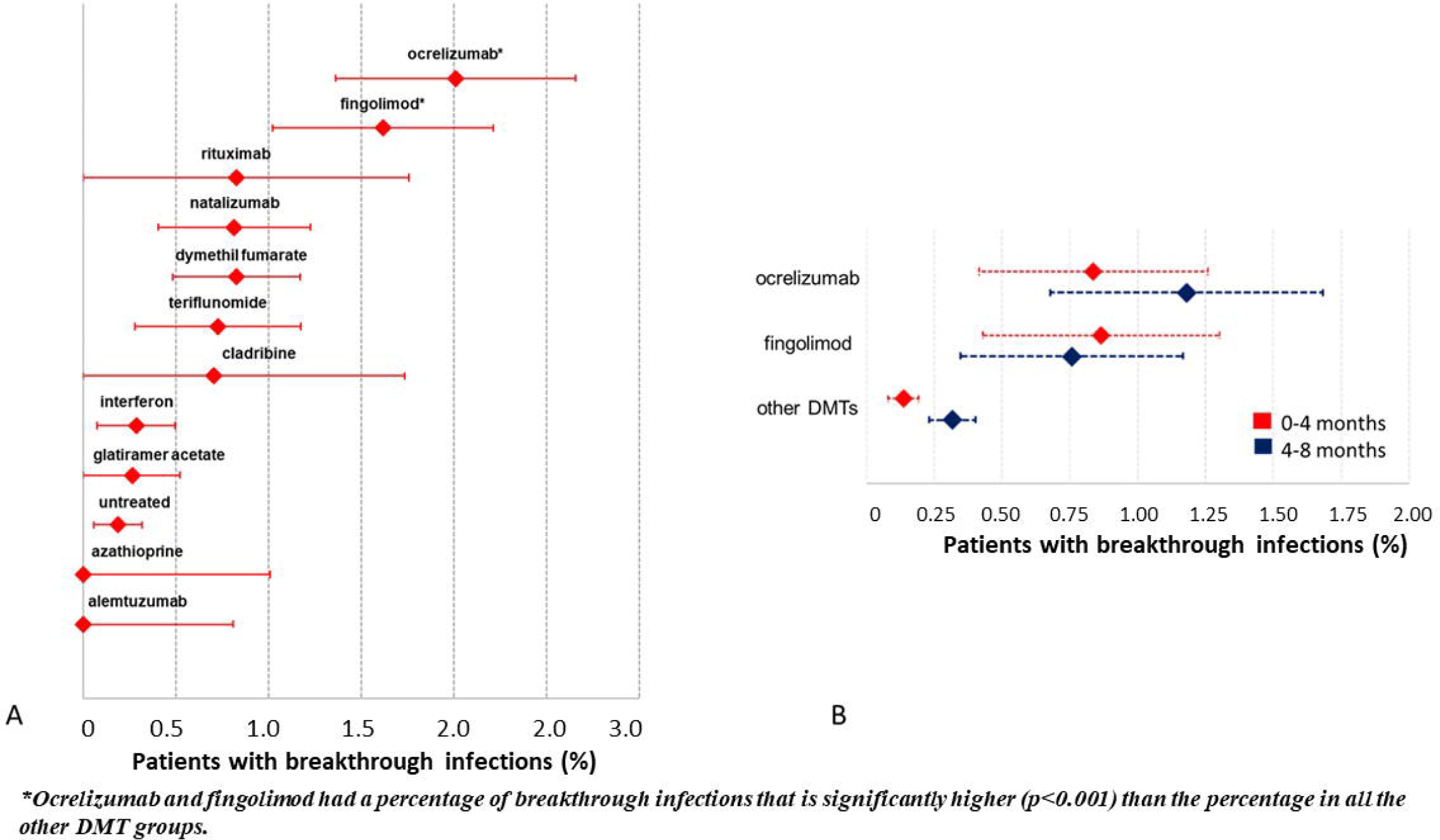
**Cumulative incidence of breakthrough infections in patients in each DMT group (A) and breakthrough infection rates according to time since vaccination in ocrelizumab, fingolimod and other DMTs (B).**

Figure1, panel B, reports the rate of breakthrough infections in two time periods of equal duration: the first 4 months following the last vaccination dose vs the period 4-8 months after the last vaccination dose in patients treated with ocrelizumab, fingolimod and all the other DMTs. The rate in patients treated with ocrelizumab and fingolimod was not significantly affected by the time since vaccination (ocrelizumab: 0-4 months after vaccination: 0.84%, 4-8 months after vaccination: 1.18%, OR=1.40, p=0.31; fingolimod: 0-4 months after vaccination: 0.86%, 4-8 months after vaccination: 0.76%, OR=0.88, p=0.75). In all the other DMT groups the rate is much lower (0-4 months after vaccination: 0.14%) and it was significantly increased after 4 months from the last vaccine dose (4-8 months after vaccination: 0.32%, OR=2.32, 95%CI=1.38-4.01, p<0.001).

## Discussion

This study on a large sample of pwMS who received a full vaccination cycle confirms that the risk of contracting SARS-CoV-2 infection after Covid-19 m-RNA vaccines is higher in pwMS on anti-CD20 monoclonal antibodies or fingolimod. We observed just one admission to ICU and no deaths. Despite the small sample of 137 infections, two results emerge. First, in our cohort, among the infected patients after vaccination treated with ocrelizumab the hospitalization rate is very similar to the hospitalization rate of patients on the same treatment in the pre-vaccination era^8^, while it is reduced by 67% in pwMS in other DMTs. However, we must consider that this result can be confounded by an increased propensity of clinician to admit to hospital pwMS on ocrelizumab who develop Covid-19, because of previous studies showing that these patients are at a higher risk for a severe course^8^. Second, as expected^9^, the vaccine-induced protection from the disease is waning with time since vaccination, and this is more evident in patients treated with DMTs other than ocrelizumab and fingolimod, who already had low antibody levels soon after the vaccination. In fact, while the infection rate is similar in the first and in the second four months after vaccination in patients on ocrelizumab and fingolimod, and consistently higher than in patients on other DMTs, the initial protective effect is vanishing with time for patients in the other DMTs group, who had a good level of antibody response four weeks after vaccination^1^. This study complements the information of previous studies reporting the antibody levels after anti-SARS-Cov-2 vaccination in pwMS on different DMTs^1-7^, suggesting that antibodies play a dominant role in preventing Covid-19 infections and their severe consequences.

## Data Availability

All data produced in the present study are available upon reasonable request to the authors

## Acknowledgements

Supported by FISM [2021/Special-Multi/001]

## Breakthrough infections in MS study group

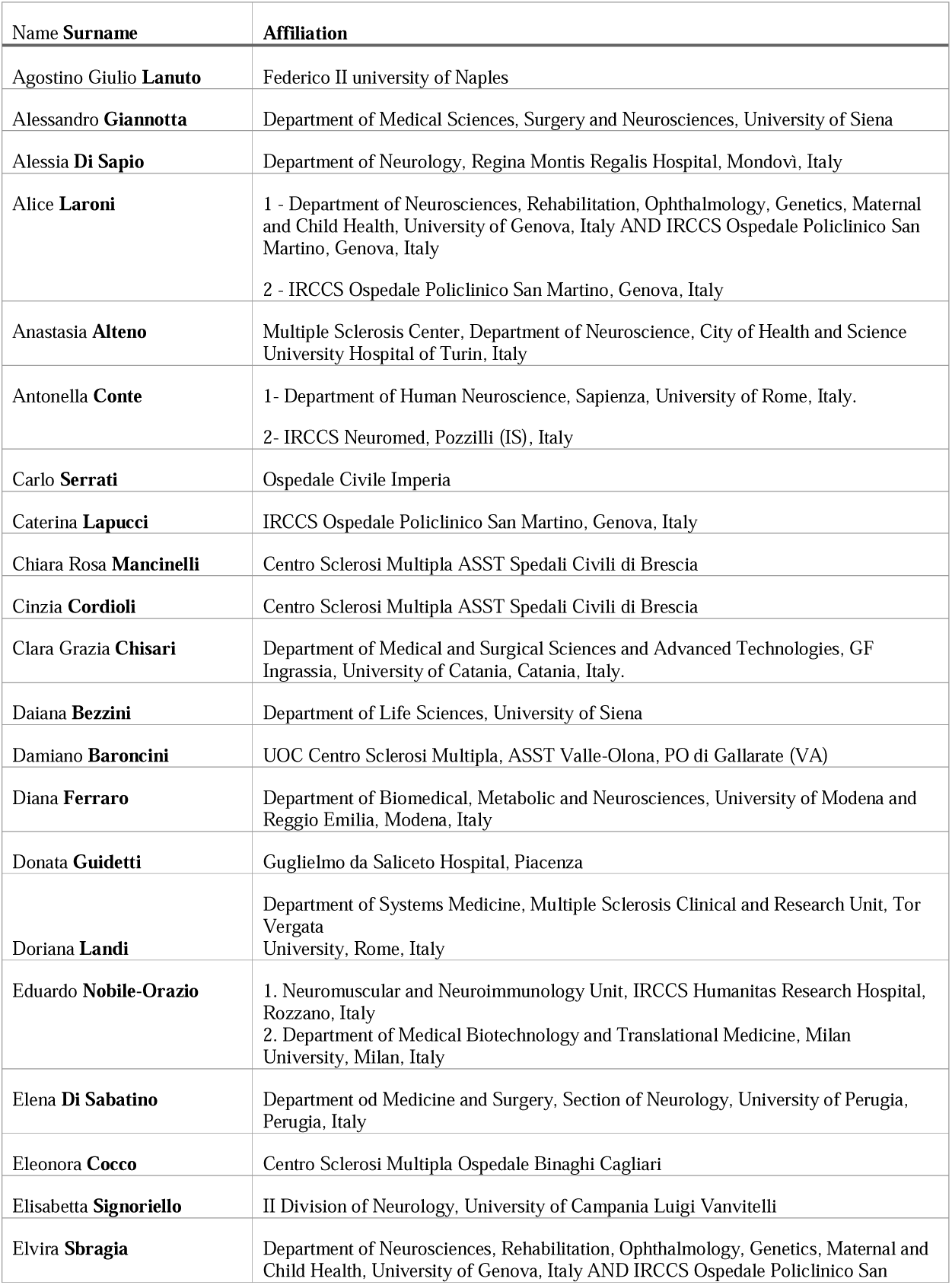
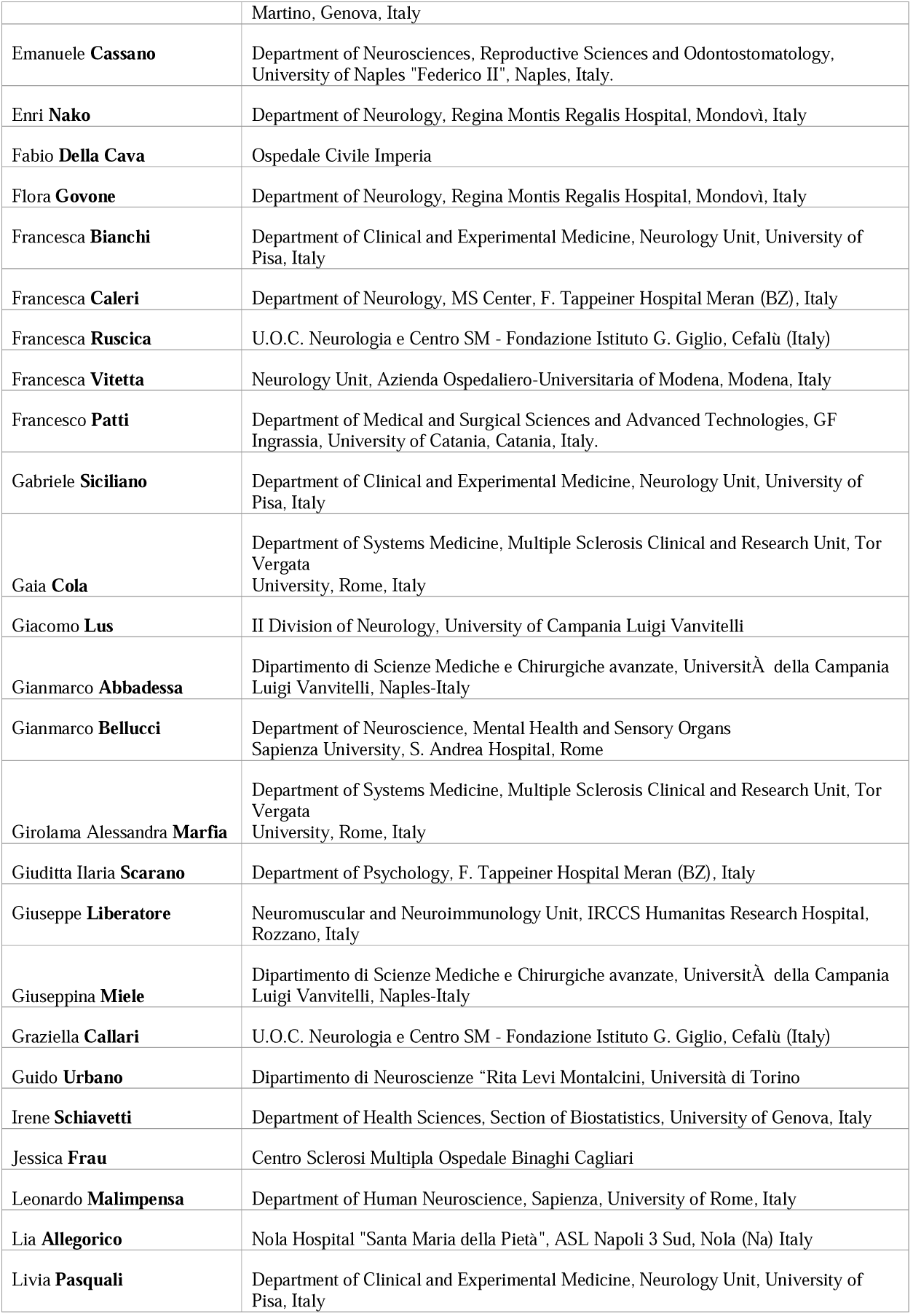
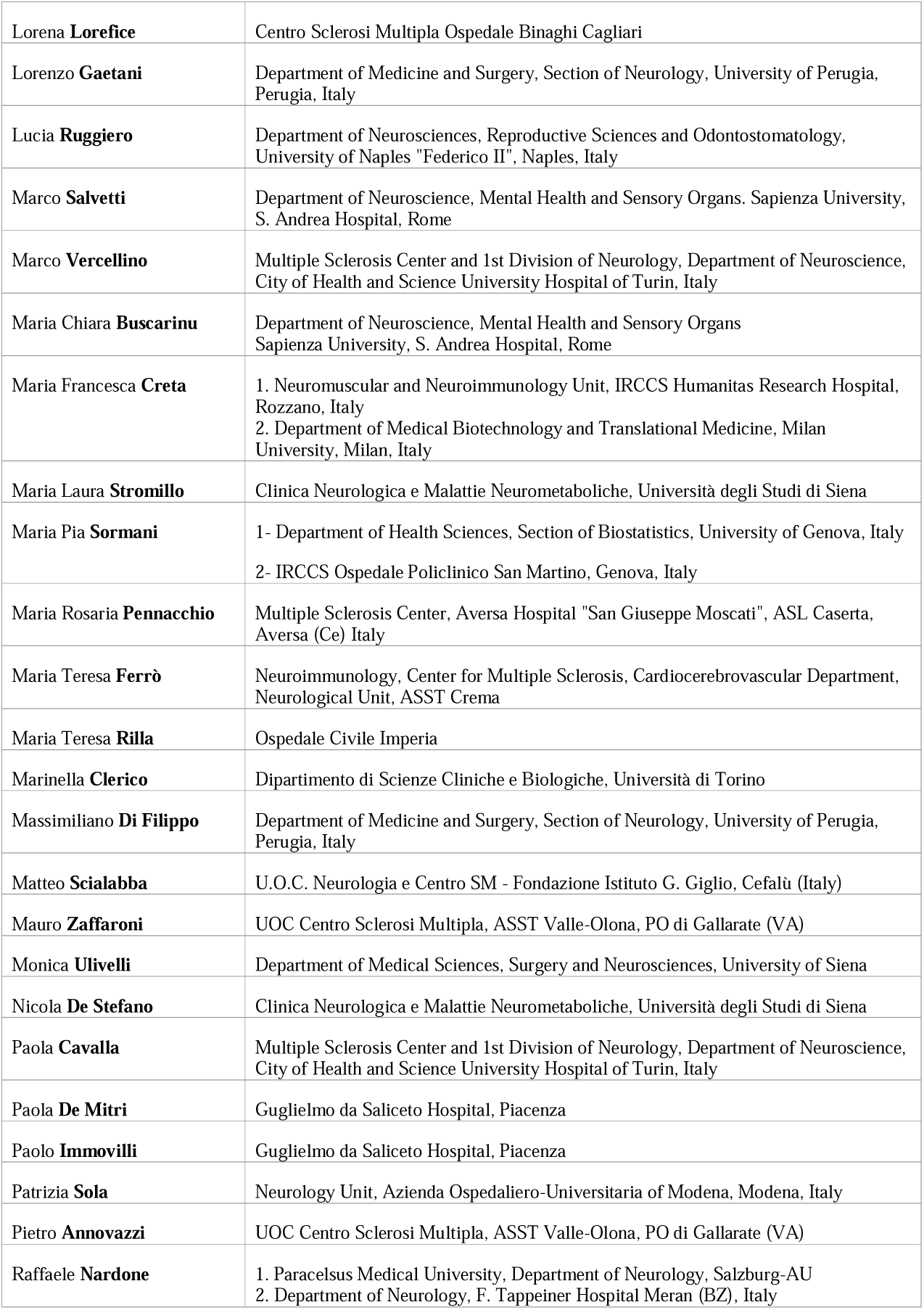
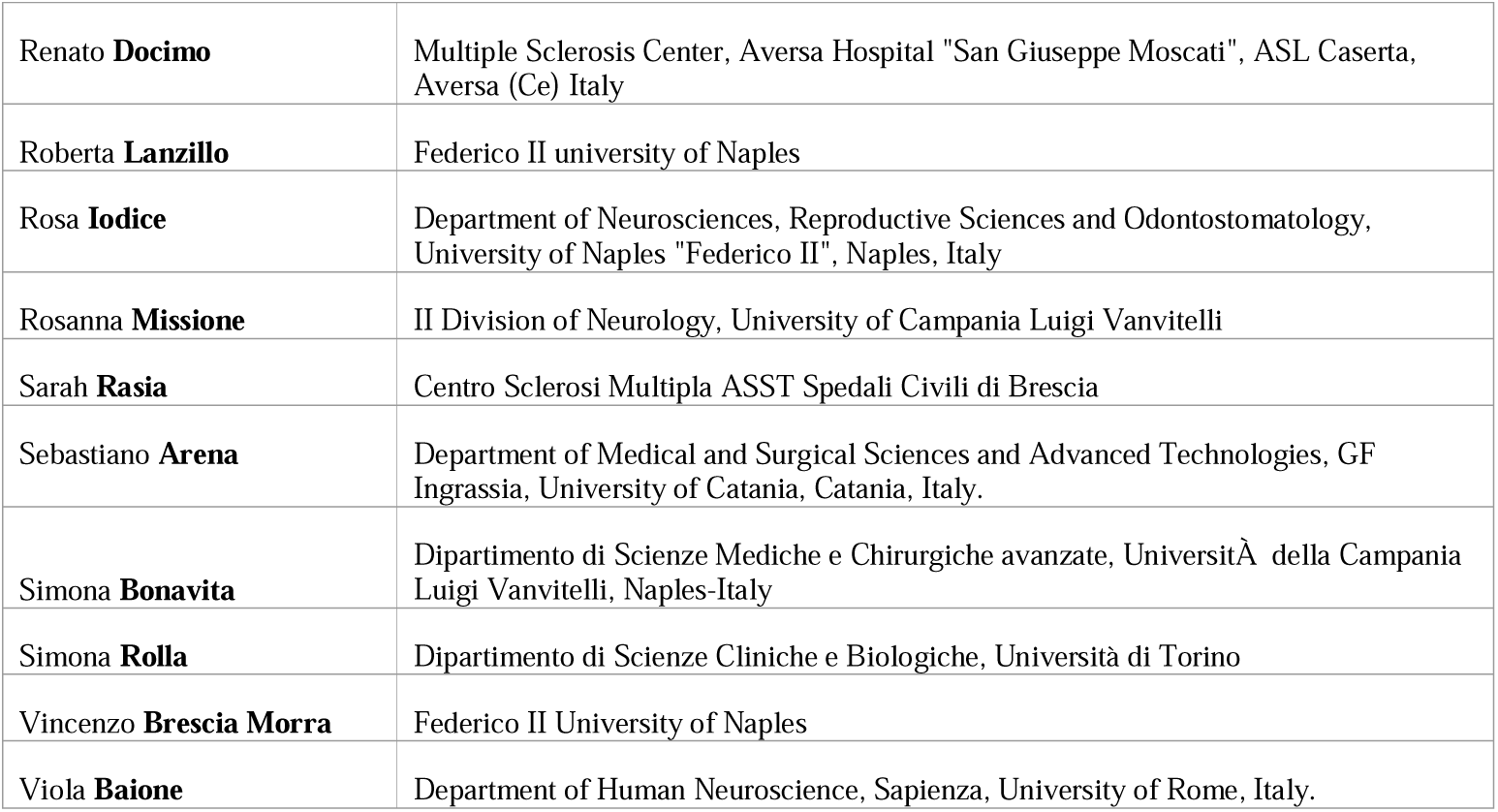

